# To test or not to test? A new behavioral epidemiology framework for COVID-19

**DOI:** 10.1101/2022.12.22.22283830

**Authors:** Jayanta Sarkar

## Abstract

Recent clinical research finds that rapid transmission of SARS-CoV-2 is facilitated by substantial undocumented asymptomatic infections. Asymptomatic infections have implications for behavioral response to voluntary testing. The paper argues that a substantial proportion of SARS-CoV-2 infections are hidden due to rational test avoidance behavior, especially among those without perceptible disease symptoms. However, if perception of disease threat is prevalence dependent, testing compliance increases in response to reported infection prevalence rate in the population. This behavior, in turn, affects infection and mortality dynamics. This paper proposes an analytical framework that explicitly incorporates prevalence-dependent testing behavior in a standard epidemiological model, generating distinctive equilibrium epidemiological outcomes with significant policy implications. Numerical simulations show that failure to consider endogenous testing behavior among asymptomatic individuals leads to over- and underestimation of infection rates at the peaks and troughs, respectively, thereby distorting the disease containment policies. The results underscore the importance of augmenting testing capacity as an effective mitigation policy for COVID-19 and similar infectious diseases.

**JEL Codes:** I12, I18

## 1. Introduction

One of the rare bright spots in the global COVID-19 pandemic is that it has offered a unique opportunity to understand the complex interactions between infectious disease transmission, human behavior, and mitigation policies. Economists have long attempted to analyze human responses to disease threats in protecting self and others, and how these responses impact disease transmission and policy. Anecdotal evidence provides many instances of voluntary protective behavior.^1^ However, the COVID-19 pandemic has given rise to ‘silent’ infections – a unique nature of disease transmission that has serious implications for preventive behavior, disease outcomes, and health policies. These silent infections spread through asymptomatic individuals who, despite being infectious, never develop perceptible symptoms. Clinical research using seroprevalence data show that the hidden asymptomatic infections account for a large proportion of COVID-19 cases and are the *main* source of dissemination of SARS-CoV-2 (Rasmussen and Popescu, 2021; Almadhi et al., 2021; Li et al., 2020). Reliable estimates based on representative sample puts the prevalence of asymptomatic cases somewhere between 40 to 45 percent of all SARS-CoV-2 infections (Oran & Topol, 2020). Asymptomatic infections, therefore, pose considerable challenges to COVID-19 mitigation efforts worldwide that are primarily reliant on symptom-driven testing strategies.

One such challenge is related to the behavior of asymptomatic individuals around testing for infection. Despite increasing vaccination rates worldwide, diagnostic testing remains the mainstay of COVID-19 management strategy in most places. While testing is mandatory in some settings (e.g., for health care workers), voluntary testing relies on human compliance. However, there are clear disincentives to testing – mainly due to the serious implications of the consequences of testing ‘positive’, which can have adverse repercussions on employment, children and family, and involve direct, indirect, and psychological cost of quarantine. These consequences are likely to be perceived more acutely by the asymptomatic agents, mostly in absence of a perceivable signal of having contracted the disease. Not surprisingly, non-compliance and refusal to undergo testing even when symptomatic has emerged as a public health problem in some countries (McDermott et al., 2020). The self-isolation or quarantine requirement following a potentially positive result, and consequent loss of income have been identified as key impediments to testing in the US and elsewhere (Rubin, 2020; Egelko et al., 2020; Thappa & Rana, 2020, Morris, 2020). A community surveillance system reported that only about 55 percent of people with fever and cough tested for COVID-19 in the week following the second peak of the pandemic in Australia in early August 2020.^2^ Given that the number of people who have tested and isolated are key inputs to the quantification of disease risk, non-compliance to testing and isolation (henceforth T&I) constitutes a significant behavioral barrier to effective infection control policies. Due to this diagnostic barrier, the true incidence of SARS-CoV-2 infection remains uncertain in many countries due to this diagnostic barrier, despite unprecedented levels of tracking and testing.

The paper conceptualizes an economic-epidemiological framework that integrates adaptive T&I behavior among asymptomatic individuals (henceforth, the *asymptomatics*) with a standard epidemiological model. In this framework, undertaking T&I is an economic decision since it involves high uninsurable private cost of lengthy self-isolation in the event of a positive test result. Thus, the absence of underlying symptoms reduces the perceived risk of being infected, diminishing the incentives for T&I among the asymptomatics. However, this perceived risk and T&I incentives rise in response to external indicators of disease threat – such as, reported infection prevalence.^3^ The ‘behavioral’ model predicts significantly higher equilibrium transmission rate and mortality compared to the standard ‘exogenous behavior’ models, thereby providing a theoretical foundation for the hidden infections and the associated ‘excess deaths’ reported worldwide.^4^ One of the key insights is that the peak of infection is self-limiting because preventive T&I behavior responds to rise in infection prevalence. On the downside, hidden infection is also predicted to linger at the tails when T&I falls with decreasing prevalence, making complete eradication of the virus extremely difficult. The simulated model successfully replicates the fast-changing daily data from Italy, which establishes the external validity of the model.

The paper contributes to the burgeoning economic-epidemiology literature by highlighting the role of economic behavior in the propagation of SARS-CoV-2. Some models incorporate mechanisms whereby disease risk simultaneously affects and are affected by individual behavior in the form of prophylactic behavior and vaccination, particularly in the context of sexually transmitted diseases, such as HIV/AIDS (Geoffard and Philipson, 1996; Hyman and Li, 1997; Phillipson, 1999, Klein et al., 2007; Kremer, 1996; Auld, 2003; Valle et al., 2005). However, endogenous behavioral responses in epidemiological models remain largely unexplored, except in a handful of studies (Klein et al., 2007; Ferguson, 2007; Carpenter, 2014; Funk et al., 2015). Eichenbaum et al. (2021) study the trade-off between economic and health costs when rational economic agents respond to the risk of infection by adjusting their consumption and work decisions in a standard epidemiology model. However, most COVID-19 inspired studies focussed on individual decisions to interact socially and economically (e.g., Bisin and Moro, 2022). As in this paper, these decisions are shaped by current infection rate, without internalizing the fact that these decisions affect the future infection rate. T&I decision as a relevant behavior has not been analysed in the literature, leaving the epidemiological phenomena of undercounting infection and fatality numbers unexplained. This paper is the first to do so using a conventional epidemiological model.

The results have important policy implications related to the success of disease management. First, as evident from recent trends, symptom-driven testing strategies fail to recognize the asymptomatics and therefore underestimate the true magnitude and severity of COVID-19. Second, the resulting under-ascertainment of infection risk may distort the level of stringency of restrictive policies, increasing the likelihood of waves of infection in the population, and consequently the burden of the pandemic. Third, underestimation of prevalence rate at low levels of infection has serious implications for policies targeted to achieving herd immunity and subsequently ending preventive social and travel restrictions. Emerging evidence on the newer SARS-CoV-2 variants suggests that the newer genetic mutations of the virus (e.g., the ‘Omicron’ variant), while more infectious, are likely to produce more asymptomatic infections.^5^ In this scenario, increasing testing capacity, such as development of effective rapid self-test kits, is shown to be an effective health policy measure for the containment and eventual extinction of the virus.

The rest of the paper is structured as follows. Section 2 sets the scene by briefly describing a SLIR model – a widely used benchmark epidemiological model. Section 3 outlines the economic rationale for prevalence dependent T&I behavior among the asymptomatics and analyses the equilibrium outcomes of a standard model that incorporates this behavioral foundation. The policy significance of the results is discussed in Section 4, while the results of a simulated model, highlighting the effectiveness of two alternative public policy measures, are elaborated in Section 5 along with replication of Italian data. Section 6 concludes.

## 2. The standard SLIR model

We begin with the pioneering model of Kermack and McKendrick (1927) that provides a simple background framework of a compartmental model. Assume that at a given time an individual can belong to only one of the four compartments – ‘Susceptible’ (at risk of infection), ‘Latent’ or exposed (infected, but not yet infectious), ‘Infectious’ (transmits the disease), or ‘Recovered’ with life-long immunity. Individuals start as susceptibles and transition through these sequential phases, ending their infection lifecycle as recovered or ‘removed’ if the disease is fatal. Let at any time *t* the number of people in susceptible, exposed, infectious, and recovered/removed compartments be denoted by *S*(*t*), *L*(*t*), *I*(*t*), *R*(*t*), respectively. Transmission of the disease follows the principle of mass action or bilinear incidence, implying that *βS*(*t*)*I*(*t*) number of susceptible individuals move from *S* to *L* group per period, where *β* is the infection transmission rate. Let *κ* > 0 be the rate at which each exposed person with latent infection transitions to the infectious stage, which implies an average incubation period of 1/*κ* days. Also, *ω* > 0 denotes the rate of recovery, indicating a mean infectious period of 1/*ω*. In a closed system with non-fatal disease, the population size is constant and is given by *N* = *S*(*t*) + *L*(*t*) + *I*(*t*) + *R*(*t*).^6^ The system of ordinary differential equations for the SLIR model is given by:

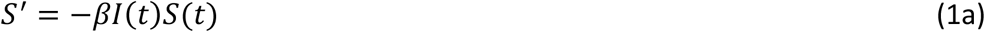

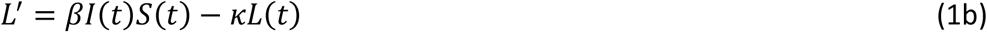

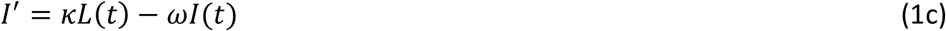

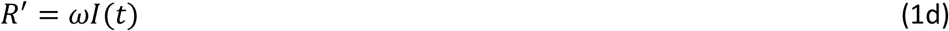

To move to a disease-free equilibrium (DFE), the number of infectious must decline with time. Evaluating the condition *L*′ + *I*′ < 0 at *t* = 0, would yield *βS*(0) < *γ*, or

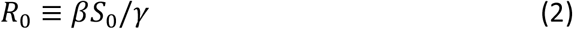

*R*_0_ is a threshold quantity, known as the *basic reproduction number*, that determines the average number of susceptible individuals a newly introduced infected person would infect in a fully susceptible population of size *S*(0) = *S*_0_. If *R*_0_ < 1, *I*(*t*) decreases monotonically to 0; whereas if *R*_0_ > 1, this number first increases, reaches a peak and declines to zero; thus *R*_0_ = 1 acts as a sharp threshold between the disease dying out or causing an epidemic. The model can generate some useful quantities, such as the maximum infection rate, fraction of population infected (attack ratio), etc. that are useful for disease containment policy.

## 3. Asymptomatic COVID-19 infections and rational disease dynamics

The standard SLIR model assumes all infection status is *known* through identifiable symptoms after a pre-symptomatic period. However, in the ongoing pandemic a significant proportion of infected individuals are known to remain asymptomatic throughout their infection lifecycle – a unique epidemiological feature of SARS-CoV-2 that is different from the preceding coronavirus outbreaks of SARS-CoV and MERS-CoV (Li et al., 2020).

The key idea explored here is that in absence of signals from physical symptoms, the asymptomatics discount the likelihood of contracting the virus and therefore have lower incentive to undertake preventive steps, such as volunteering to T&I. The implications of testing positive may deter them from testing. If tested positive, they will be unable to leave their homes, return to work, and spend time with their families for a considerable time. These private costs are uninsurable but can be avoided by not opting to test. There is some evidence that people with asymptomatic or mildly symptomatic infections typically fail to present at healthcare facilities or appear for testing (Kim et al., 2020; Nishiura et al., 2020). Some refuse to test, even when symptomatic (Contreras, et al., 2021). Lack of incentive for T&I may be more widespread among the asymptomatics, resulting in many undetected transmissions. Indeed, recent scientific evidence strongly suggests that these hidden infections are the driving force behind the surge of infections of SARS-CoV-2 (Rasmussen & Popescu, 2021; Almadhi et al., 2021; Li et al., 2020).

Nonetheless, maladaptive behavior is amenable to change with infection prevalence – an idea explored in Geoffard and Philipson (1996), Auld (1997), Philipson (1999) where demand for prevention – specifically, through vaccination – rises in response to infection rate. Results from a recent randomized controlled trial in Mozambique demonstrate that adherence to preventive policies, such as social distancing, responds positively to rates of infection (Allen et al., 2021). In the COVID-19 context, T&I is the first step in preventing unintentional community transmissions. Hence, in absence of signals from physical symptoms, private demand for T&I among the asymptomatics is assumed to depend upon current infection prevalence rate.^7^

### 3.1 An augmented SLIR model with rational preventive behavior

Let us set the background by providing a theoretical argument for prevalence dependence of T&I behavior. Here, the infected individuals with no or little symptoms are unaware of their infectivity and the threat they pose to others. Hence, their T&I decision is informed by the perceived probability of being exposed to the virus and the associated costs and benefits. Let *β* ∈ (0,1) be the transmission probability. If the probability of a random contact occurring between a susceptible and an infected (following the principle of *mass action*) be given by the prevalence rate *i* ≡ *I*/*N*, the probability of an infected person infecting a susceptible at any time *t* is given by *p*_*t*_ = *βi*_*t*_. This formulation is similar to Geoffard and Philipson (1996), Philipson (1999) and Jones et al. (2020) where the probability of contracting an infectious disease by a susceptible is proportional to the existing prevalence rate.

Let 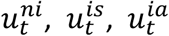 denote the utilities or payoffs of a non-infected, infected with symptoms (symptomatic), and infected without symptoms (asymptomatic) agent from own health at any time *t*, respectively. Disease symptoms, however mild, reduce utility from health so that 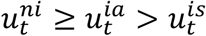. In addition to their own health, individuals also derive utility from health of the susceptible people they live, work, and socialize with (close contacts), collectively given by 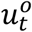, and let *φ* > 0 denote the importance of this social utility. An infected agent transmits the virus to a susceptible close contact with probability *p*_*t*_, which reduces *u*^*o*^ by a factor *z* > 0. Assume that individuals incur a positive cost *c*_*T*_ > 0 if they want to get tested, which may depend on transport cost to the testing centers, availability of testing services, psychic and physical discomfort from the test procedure, etc. For simplicity, tests are assumed to be perfectly accurate. Conditional on testing positive, an infected must self-isolate or quarantine for a considerable period (generally 14 days), and in the process incurs a cost: 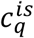 and 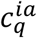 for the symptomatics and the asymptomatics, respectively. Because the psychic cost of quarantining is likely to be higher for the asymptomatics we expect 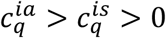. For the expositional ease, let *ϑ* > 0 be the probability of death for an infected, regardless of symptoms, and *α* > 0 be the temporal discount factor.

The decision to test is analyzed simply in a two-period model framework. Infected individuals are either symptomatic or asymptomatic, who seek to maximize the expected utility over the two periods by choosing whether to undergo testing. In period *t* an agent is aware of possible exposure to the virus, considering the option of testing in period *t*+1, given symptoms or absence thereof. To do so, she compares the expected utilities corresponding to the outcomes with and without testing. A symptomatic person, whose current utility is 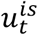 weighs the ex-ante utilities from testing vis-à-vis not testing, taking the probability of being infected, *p*_*t*_ to be given. All future utilities are weighted by the effective discount factor *α*(1 − *ϑ*). If she decides not to test and survives the next period, her future utility from own health is 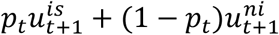 and that from the health of close contacts is 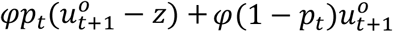. If she undergoes testing and isolation, she incurs a current cost *c*_*T*_, and receives a future utility of 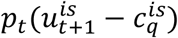. Similarly for the asymptomatic individuals. Let the expected utility of a symptomatic infected agent is given by:

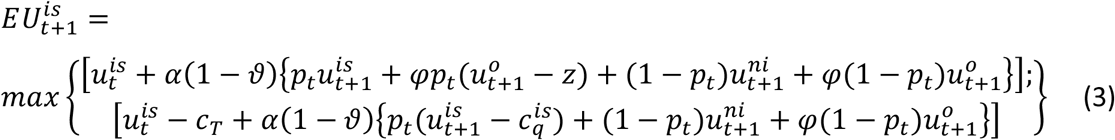

Similarly, the expected utility of an asymptomatic agent is:

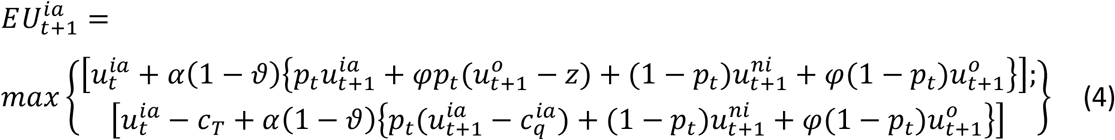

It can be shown that there exist uniquely continuous expected utility functions that satisfy equations (3) and (4). From equation (3), a symptomatic individual will choose to test *iff*:

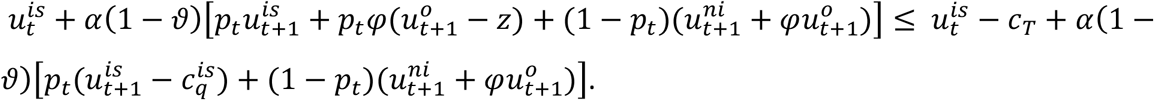

This condition simplifies to yield a threshold probability of testing for the symptomatics 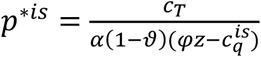, and for the asymptomatics, 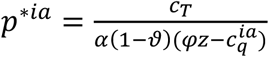. Given *p* = *βi*, these quantities also reflect the critical disease prevalence rate that triggers T&I behavior. The threshold probabilities increase in the cost of testing and that of isolation and decrease in the cost of health of close contacts. Thus, the higher the T&I-related costs and lower one’s level of care for others’ health, an infected agent would postpone testing until infection prevalence increases further. Given 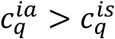, this threshold is higher for the asymptomatics, who are expected to hold off testing further than the symptomatics, given everything else.

Distinct from the existing studies on prevalence-dependence, the perceived exposure probability and hence the probability of T&I is assumed to depend on the *reported* prevalence rate, which is based on the number of detected positive cases, *T*(*t*). In what follows, the probability of undertaking T&I by a symptomatic and an asymptomatic would be given by *p*^*s*^ and *p*^*a*^, respectively, where *p*^*a*^ is increasing in infection prevalence rate.

To account for the behavioral feature described above, I augment the canonical SLIR model by including a latent or gestation stage and a T&I stage preceding the Recovery state. The latent stage allows for some time for the state of infection to be revealed as symptomatic or asymptomatic. We assume a fraction ϕ of the infected population becomes symptomatic. The T&I stage *T*(*t*) captures the part of the population who choose to test and isolate at time *t*. For simplicity, all tests for infected individuals are assumed to return positive results with perfect accuracy and therefore these individuals undergo quarantine or self-isolation with full compliance. Thus, people in the *T* compartment are non-transmitter of the disease. The transition across health states in the proposed Susceptible-Latent-Infectious-Tested/Isolated-Recovered-Dead (SLIITReD) model can be represented in Figure 1.

**Figure 1:**
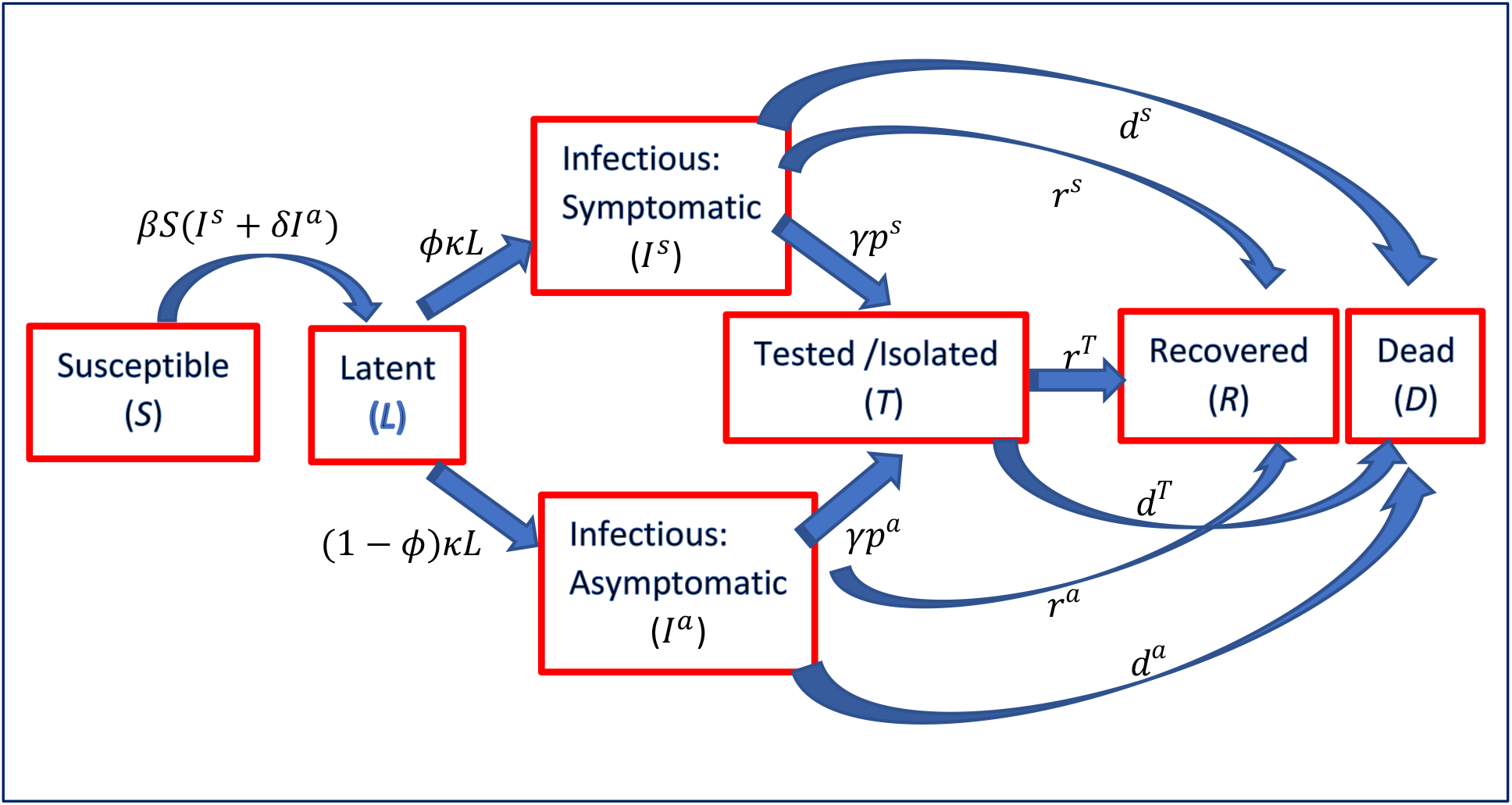
Disease transmission in the SLIITReD model

Given that the information set for the observed epidemiological states is denoted by {*S*(*t*), *T*(*t*), *R*(*t*), *D*(*t*)}, the asymptomatic individuals’ beliefs that they have indeed been exposed to the virus would be proportional to the reported positive cases *T* as a fraction of people in these epidemiological states (either susceptible or known to have been infected). Thus, the probability that the asymptomatics assign to being exposed, hence their probability of undertaking T&I at time *t*, is given by *p*^*a*^(*t*) = *μT*(*t*)/[*S*(*t*) + *T*(*t*) + *R*(*t*) + *D*(*t*)], where *μ* > 0 is a sensitivity parameter.^8^ Given *p*^*a*^(*t*) *≤* 1, *μ ≤* max [{*S*(*t*) + *R*(*t*) + *D*(*t*)}/*T*(*t*)]. Note that *p*^*a*^(0) = *μT*_0_/(*S*_0_ + *T*_0_), where *S*(0) = *S*_0_ > 0, *T*(0) = *T*_0_ ≥ 0 and 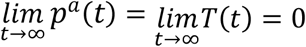. Normalizing *N* = 1, the dynamic system the model is represented by the following ODEs:

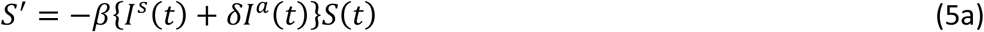

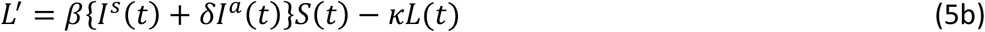

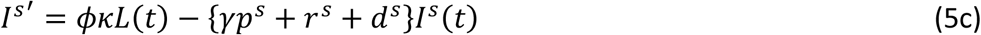

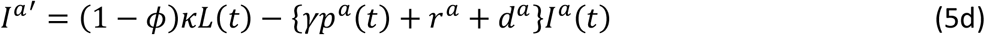

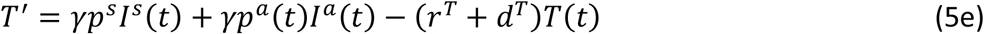

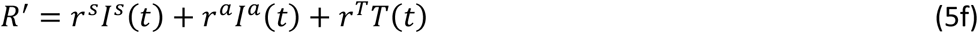

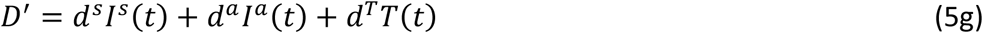

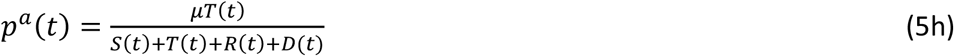

As in the mainstream epidemiological models, the incidence rate is assumed to be bilinear for simplicity. The asymptomatics are assumed to carry a lower viral load and therefore have a lower infectivity denoted by *δ* < 1. Average latency period is 1/*κ*; the death rate and recovery rate of the symptomatic infected are *d*^*s*^ and *r*^*s*^, while these values for the asymptomatics are *d*^*a*^ and *r*^*a*^, respectively. The recovery and death rates under isolation is *r*^*T*^ and *d*^*T*^, respectively.

This system nests the standard SLIR model which can be recovered by setting *I*^*s*^(*t*) = *I*^*a*^(*t*) and *T*(*t*) = 0; *p*^*s*^ = *p*^*a*^ = 1; *r*^*s*^ = *r*^*a*^ = *r*; *d*^*s*^ = *d*^*a*^ = 0. As in the standard model this behavioral model describes diseases in which recovered individuals are immune to reinfection. Note that the Mass conservation property holds, because *S*′ + *L*′ + *I*^*s*^′+ *I*^*a*^′ + *T*′ + *R*′ + *D*′ = 0. Hence, *S*(*t*) + *L*(*t*) + *I*^*s*^(*t*) + *I*^*a*^(*t*) + *T*(*t*) + *R*(*t*) + *D*(*t*) = *N* = 1. The system with seven differential equations is positive as all state variables take non-negative values for *t* ≥ 0, given non-negative initial conditions. Note that *R*(*t*) and *D*(*t*) are cumulative variables that depend only on the other ones and their own initial conditions.

Given the set of initial conditions {*S*(0), *L*(0), *I*^*s*^(0), *I*^*a*^(0), *T*(0), *R*(0), *D*(0)} and *p*^*a*^(0) ≥ 0, the variables converge to the *DFE* given by: 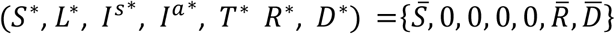, with 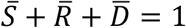. This set represents all possible equilibria. Appendix A derives the basic reproduction number using the new-generation approach as the maximum element in the spectrum of the next-generation matrix, as in the modern mathematical epidemiological models. The behavioral *R*_0_ is given by:

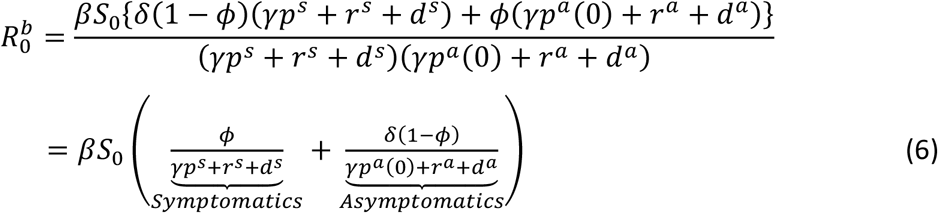

Thus, 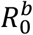 neatly decomposes into two parts: 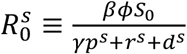 and 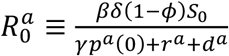, such that 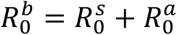. Standard predictions that fully discount the existence of asymptomatic infections, would severely underestimate *R*_0_ by omitting the second term in equation (6). Furthermore, given 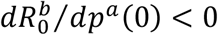, the standard models that do recognize asymptomatic infections but assume exogenous and uniform T&I behavior across the infective groups *p*^*a*^(0) ≈ *p*^*s*^, would also underestimate *R*_0_.

As expected, more effective epidemic containment measures, such as social-distancing and lockdowns, that reduce the magnitude of *β* reduce 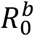. Focusing on 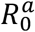, note that *βδS*_0_ are the newly exposed individuals generated by one asymptomatic infectious individual per unit of time in an entirely susceptible population. A fraction 1 − *ϕ* of them progress from the exposed stage to the infectious stage, each of whom remains infectious in the community for 1/(*γp*^*a*^(0) + *r*^*a*^ + *d*^*a*^) periods. Therefore, 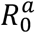 denotes the number of secondary infections that one asymptomatic will produce in an entirely susceptible population during his/her lifespan. 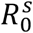 denotes the counterpart for a symptomatic.

As always, if 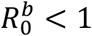 there will be no epidemic. An epidemic will ensue in a virgin population if 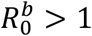. In the latter case, infection cases will increase, but will eventually fall because *S* falls monotonically (*S*’ < 0 for all *t*). Denoting 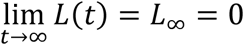 and 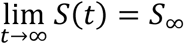, the final size relation is given by: 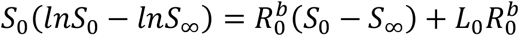. The incidence rate among the symptomatics is *ϕ*(*S*_0_ − *S*_∞_)/*S*_0_ and that among the asymptomatics is (1 − *ϕ*)(*S*_0_ − *S*_∞_)/*S*_0_.

Once the outbreak is underway, the interaction between testing behavior and prevalence makes the reproduction number time varying. The e*ffective* reproduction number at any time *t* > 0, is given by:

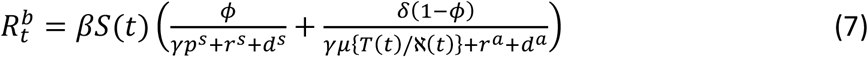

where, ℵ(*t*) ≡ *S*(*t*) + *T*(*t*) + *R*(*t*) + *D*(*t*). Higher testing rate (*γ*) and greater rates of transition from the infectious category in the forms of increased recovery and death rates decrease 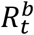. The innovation in this behavioral model is that the effective reproduction number depends on the endogenous probability *p*^*a*^ through the quantity *T*(*t*)/ℵ(*t*). Specifically, the magnitude of 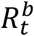 is decreasing in *T*(*t*)/ℵ(*t*) – meaning, given everything else, the higher this ratio, the higher is the probability of T&I for an asymptomatic, the faster is the rate at which the infectious are removed from future contact possibilities with the susceptibles. Therefore, a greater sensitivity of testing behavior to infection prevalence among the asymptomatics (*μ*) decreases both 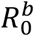 and 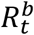.

Given the critical role of adaptive T&I behavior, it is useful to know the magnitude of *p*^*a*^ that determines the threshold around which infections change course. This threshold can be derived by solving the value of *p*^*a*^ at which infection rate is maximum – that is when *I*^*s*^′ + *I*^*a*^′ = 0, which obtains when the rate at which infectious people enter, *κ*, equals the rate at which they are removed either by isolation, recovery or death *ϕ*(*γp*^*s*^ + *r*^*s*^ + *d*^*s*^) + (1 − *ϕ*)(*p*^*a*^(*t*) + *r*^*a*^ + *d*^*a*^). This condition solves 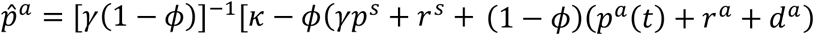. Therefore, infection rate rises as long as 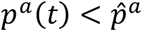, and falls otherwise. The magnitude of 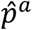 can be inferred from data.

## 4. Significance for public health policy

The behavioral model has serious implications for the reliability of SARS-CoV-2 fatality measures such as Case Fatality Rate (CFR) and Infection fatality Rate (IFR). The CFR is typically defined as the number of deaths from COVID-19 as a proportion of the number of people who tested positive for the virus, while IFR is defined as the proportion of death among all infected people. However, the behavioral model highlights the undetected deaths and infections are a significant source of uncertainty for these observed fatality rates. First, given that the mortality among the asymptomatics is similar to that of the symptomatics, COVID-19 deaths from undetected infections are likely to be significant (Park et al. 2021).^9^ Second, the significant number of undetected asymptomatic cases leads to substantial underestimation of the true case numbers.

Given the testing rate *γ* and T&I probabilities, the true CFR at time *t* is defined as 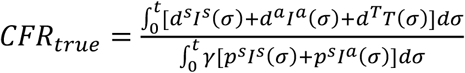, which obtains when no differences in T&I compliance exist between the two infective groups – that is when they share the same probability of testing, *p*^*s*^. The reported CFR, however, depends on actual diagnostic probability *p*^*a*^(*t*) and the associated undetected fatalities *d*^*a*^*I*^*a*^(*t*), and can be represented as 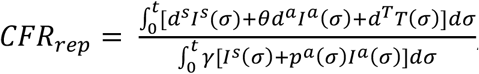, where *θ* ∈ [0,1] is the fraction of deaths among the undiagnosed asymptomatics that are eventually (and correctly) attributed to COVID-19. At low rates of infection prevalence, *p*^*a*^(*t*) is low, although the detected COVID-19 deaths are likely to be high. Thus, *CFR*_*rep*_ is likely to exceed *CFR*_*true*_. Eventually, *CFR*_*rep*_ decreases with rising prevalence rate as *p*^*a*^ rises in response. At high prevalence rates, *CFR*_*rep*_ is potentially lower than *CFR*_*true*_ if the detection rate exceeds the rate of undetected deaths, that is *p*^*a*^ > *θd*^*a*^. Figure B1 in the Appendix illustrates the extreme cases of *θ* = 0 and *θ* = 1. Similar uncertainties and biases afflict the IFR measure. In the absence of population-wide serological testings to determine prior infection status, these fatality measures are rendered unreliable. Consequently, any policy decisions based on these numbers would be flawed.

In contrast to the standard epidemiological models, growth of infectious disease predicted by the behavioral model is self-limiting. This occurs because infection prevalence induces T&I behavior among the asymptomatics, reducing the pool of infectives, and preventing sharp spikes in infection. However, containment of a disease becomes increasingly more challenging with declining disease prevalence and falling likelihood of T&I. This implication is critical for containment policies that are primarily based on T&I behavior. Similar inferences can be drawn from vaccine-based prevention policies when demand for is prevalence dependent (Philipson, 1999).

The behavioral model offers critical insights for restrictive epidemic management policies, such as lockdown. Interestingly, the optimality of stringency of lockdown depends on whether stringency is determined by the effective reproduction number, or by the simpler measures such as infection and/or fatality rates. To elucidate the difference in policy biases, let us analyze a social planner problem who can lockdown a fraction *π*(*t*) ∈ [0, 1] of the population. First, let the stringency of lockdown, *φ*_*t*_ ∈ [0,1], depends positively on the effective reproduction rate – i.e., 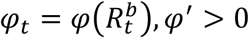. Individuals are assumed to die only from infection. In absence of a lockdown, each individual produces *y* units of output. If $*V* denotes the value of a statistical life, the planner should decide to lockdown if discounted cost of output foregone due to lockdown at the desired level of stringency is less than the discounted value of lives lost in absence of it – i.e., if 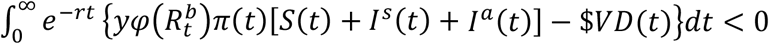, where *r* > 0 is the planner’s discount rate. Since *p*^*a*^ is overestimated and the magnitude of 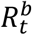 underestimated at low values of infection, the desired stringency level 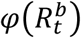 is likely to be *underestimated* in the initial and late phases of the pandemic. Hence, given everything else, the cost of death from less stringent lockdowns is likely to be suboptimally high at low levels of prevalence.

On the other hand, if the level of stringency is determined by the detected infection rate *T*(*t*)/*S*(*t*) that significantly discounts the number of silent infections, the level of required stringency *φ*(*T*(*t*)/*S*(*t*)) would always be *underestimated* in the initial and late phases of the pandemic when *p*^*a*^ is low. This would imply a completely orthogonal pandemic management policy relative to the case above.

Given the policy dilemma regarding lockdown stringency stems from the uncertainty around *p*^*a*^, it is worthwhile to discuss the utility of non-pharmaceutical policies such as increasing (i) testing capacity (testing rate, *γ*) and (ii) testing compliance through a targeted public health campaign that aims to increase the responsiveness of *p*^*a*^ to observed infection rate, *μ*. Increasing testing capacity is equivalent to a reduction in actual cost testing – for example, through promotion of the use of effective rapid self-test kits (e.g., rapid antigen test). Increasing testing compliance through sensitizing the infected group is likely to reduce the psychic cost of self-isolation highlighted in section 3. In what follows, a parameterized model is simulated to explore the differential predictions of the behavioral SLIITReD model and a baseline ‘standard’ model that incorporates asymptomatic infections but assumes an exogenous T&I behavior. Next, the simulated model analyzes the relative epidemiological effects of the two policies that relate to increasing testing capacity and testing compliance.

## 5. Predictions of the Behavioral model

The parameter values reported in the literature are marked by substantial variation in demographics, pandemic phases, and depends on non-standardized definitions of detection rate, fatality rate, infectious period, etc. Numerical determination of these parameters is a significant challenge, given that an infinite number of different parameter sets could match the data equally well (Giordano et al., 2020). However, the main purpose of this section is not to provide quantitatively accurate predictions of the SLIITReD model, but instead offer an analytical comparison of the epidemiological outcomes between the standard and the behavioral model and depict the policy implications of the latter. The quantitative validity of the behavioral model is tested by recalibrating the parameters and replicating real infection data from Italy. This exercise is relegated to Appendix B.

The infection transmission rate *β* is assigned a value of 0.9 to generate a peak infection at around the 60-day mark from the onset of the pandemic. The values for *κ* (transition rate from latency to infectious state) is 0.2, reflecting a latency period of 5 days (Baccini et al., 2021). The value of *ϕ* (proportion of symptomatic infections) is chosen to be 0.6 following the current estimates based on representative sample (Oran & Topol, 2020). Baccini et al. (2021) estimate a median period of 4 days from symptoms to test, which imply a testing rate *γ* = 0.25. The magnitude of *δ* (transmission factor for asymptomatic infections) is not well-estimated in the literature. Using Italian data Giordano et al. (2020) suggest an effective magnitude (=*βδ* in this model) for asymptomatic transmission of 0.57 on day 1 of the outbreak, implying a maximum magnitude of 0.63. A value *δ* = 0.6 is chosen. The values of *r*^*s*^ and *r*^*T*^ are based on the estimated 28-day time interval from symptoms to recovery for the undetected symptomatics (i.e., *r*^*s*^ = 1/28) and a 24-day interval from test to recovery (i.e., *r*^*T*^ = 1/24). Using German data Grimm et al. (2020) estimated a recovery time for asymptomatic infections of 10 days, implying *r*^*a*^ = 0.10. The deaths rate *d*^*T*^(death rate among the diagnosed) is assigned a value of 0.01 as in Giordano et al (2020). The death rates *d*^*s*^ and *d*^*a*^ for the undetected symptomatics and asymptomatics, respectively, are both assigned a value of 0.01 following Park et al. (2020).

Finally, the free scaling factor *μ* in *p*^*a*^ is assumed to be 0.2, implying an average magnitude of *p*^*a*^ of 4.15 percent, which is much higher than the best available estimate reported in Baccini et al. (2021). The probability of testing for the symptomatics is assumed to be 0.5 – the upper limit reported in Contreras et al. (2021), which generates a daily effective testing rate of 12.5 percent among the symptomatic population. These parameters are identical in the benchmark ‘standard’ model that identifies the two infective groups as well as the ‘Test/Isolated’ compartment (for comparability) but assumes a probability of undertaking T&I among the asymptomatics (*p*^*a*^ = 0.04) that equals the mean value of *p*^*a*^ in the SLIITReD model.

Figure 2 illustrates the evolution of some key epidemiological measures over a 200-day horizon in the SLIITReD model vis-à-vis the standard model. Given the above parameterization, all differences between the two models should be attributed to the temporal difference in the asymptomatic T&I behavior. As conjectured, the standard model underpredicts this behavior, especially as observed infections rise. Prevalence dependent behavior thus flattens the peak in detected cases as the asymptomatic ramps up their T&I behavior in response to increasing infection rate. As a result of the undetected infections, the standard model in Figure 2 grossly underpredicts the death rate and over-predicts recovery rate in equilibrium. In both model 99 percent of the population is infected at the end of the horizon. Figure 2 shows that the behavioral model generates a higher mortality rate of about 16.4 percent and a recovery rate of approximately 83.1 percent. In the standard model, these death and recovery rates are 15.9 percent and 83.7 percent, respectively. Note that, conditional on 99 percent of the population being infected, underestimation of true CFR translates to a quantitatively similar underestimation of the true IFR.

**Figure 2:**
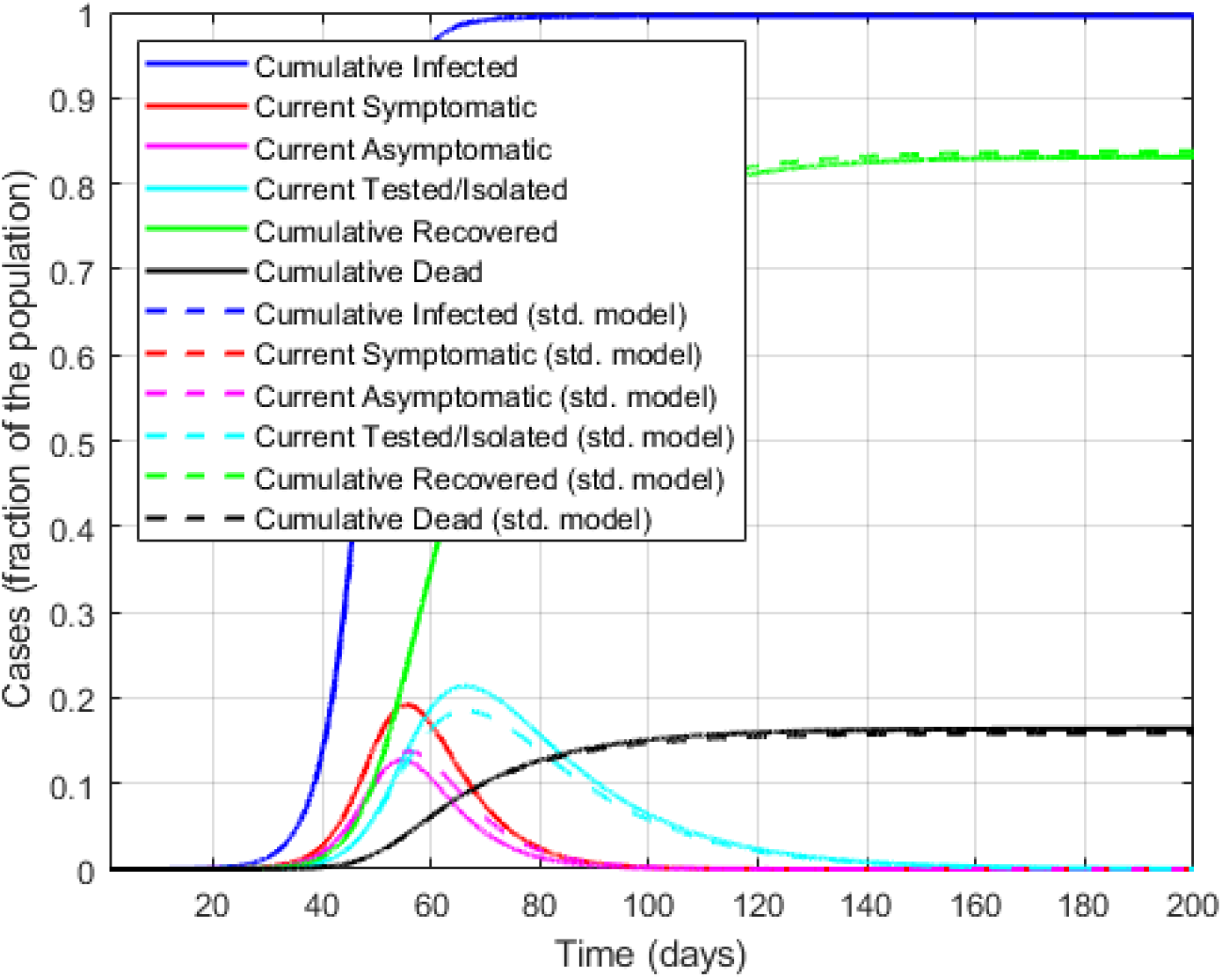
Qualitative model predictions of the SLIITEReD model versus the standard model with exogenous behavior

The implied value of the basic reproduction number (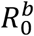) is 5.12, falling to its lowest value of 0.04 at around the 100^th^ day. The probability *p*^*a*^ also follows the infection curves with a lag, attaining a maximum value of 0.06 attained around day 65, before levelling out to 0 at around day 160. While the magnitudes of these differences are sensitive to the chosen parameters values, the basic insight is clear: the presence of undetected infections and failure to identify adaptive T&I behavior underestimate both COVID-19 fatality rate and the duration of infection prevalence.

Let us turn to examining the policy impacts of increasing (i) testing capacity (testing rate, *γ*) and (ii) testing compliance, e.g., through a targeted public health campaign. Since both policies aim to increase detection of the asymptomatic cases, the infection dynamic, recovery and death rates, as expected, depend on the magnitudes of the recovery and fatality rates among the asymptomatic cases relative to those of the diagnosed. For example, greater diagnostic efforts would lower overall death rate only if the fatality rate among the undiagnosed is higher than among the diagnosed, which is true for the chosen parameter values. Hence, the policy impacts of increasing testing compliance and testing capacity should promote testing, increase overall recovery rate and reduce overall mortality rate. We explore the effects of these two competing policy impacts in Figure 3.

**Figure 3:**
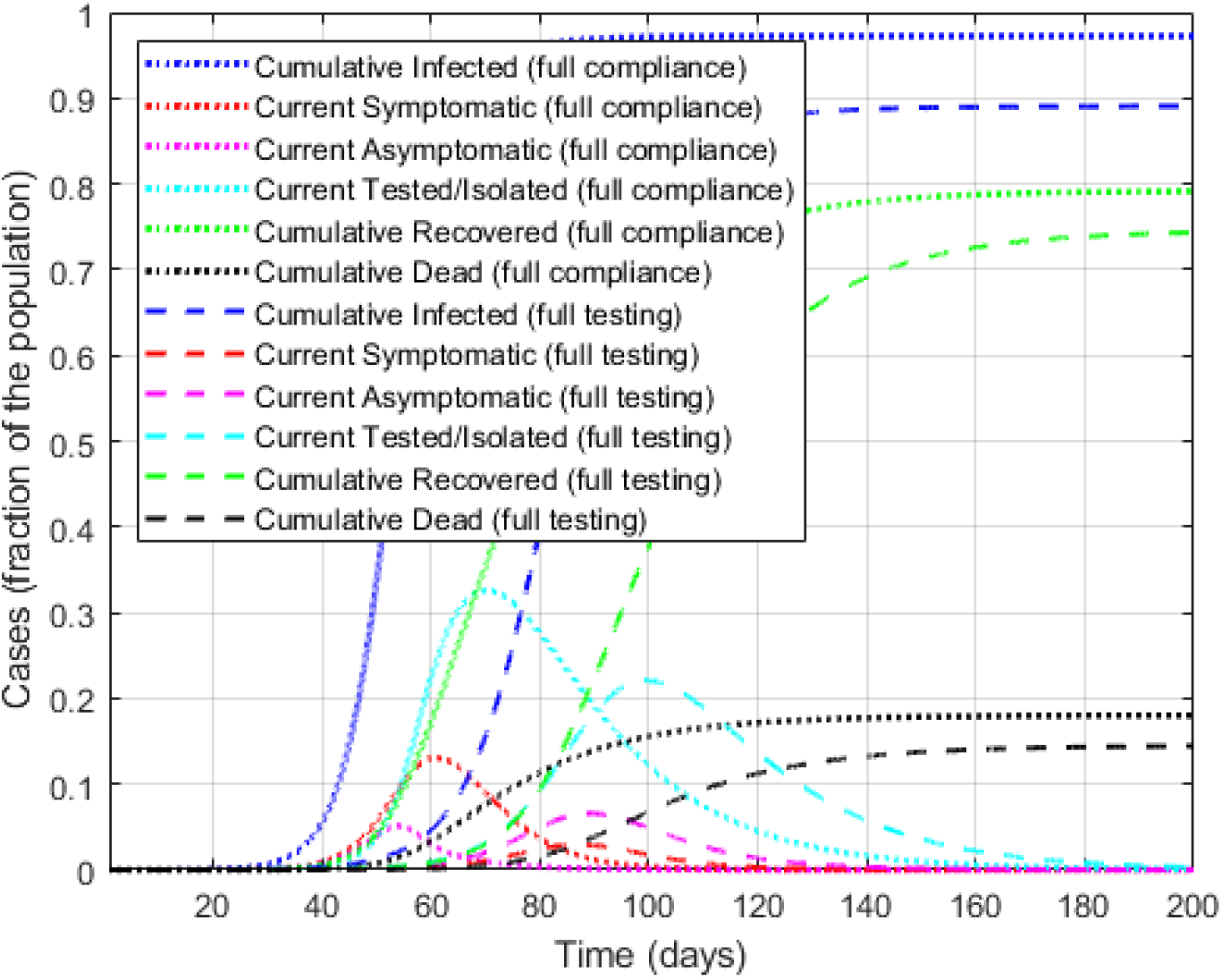
Behavioral model predictions for policies with (i) full test compliance and (ii) full testing capacity

Figure 3 compares the hypothetical scenarios under two alternative policies: a public awareness campaign that increases the probability of testing among the asymptomatics to the level of the symptomatics, and a mass testing campaign that achieves the maximum testing capacity. The awareness campaign increases the value of *μ* in *p*^*a*^ from 0.2 to 6.26 so that the *mean* of 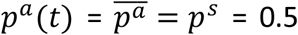, the upper limit of symptom-driven testing rate (Contreras et al. 2021) This intervention is targeted to the asymptomatic because the symptomatics already have the maximum incentive to test. Under the full testing policy, the magnitude of *γ* that reflects testing rate, increases from 0.25 to 1.

First, a comparison between Figures 2 and 3 makes it clear that relative to the baseline, both policies would ‘flatten’ the infection curves through increased testing. Counterintuitively, these interventions also lower cumulative recovery rates are increase and fatality rates. This is due to two reasons. First, overall infection rate is lower under both policies, resulting in fewer recoveries and deaths. Second, the undetected asymptomatics have a higher recovery rate (0.10) than those who test and isolate (0.042). Thus, increased testing artificially slows down recovery for the asymptomatics, which drives the fatality rate up in this compartmental model.

A comparison between the two policies in Figure 3 reveals that under the full testing policy the infection curves are much flatter and long tailed compared to awareness campaign policy. This is because the latter increases testing only among the asymptomatics, while the former allows greater testing for both infective groups, effectively spreading the testing over a longer period. The overall infection rate is much lower under full testing policy, which results in lower recovery and fatality rates. Clearly, increasing testing capacity is more effective in achieving the key outcomes of ‘flattening the curve’, overall burden of infection and lowering disease mortality, and should be a preferable disease management policy. This is echoed by the infectious disease experts in the literature.

## 6. Discussion

Worldwide, the diagnosis of COVID-19 has prioritized testing the symptomatics. The resulting proliferation of undiagnosed and undocumented asymptomatic cases induced a substantial degree of uncertainty around the true prevalence and severity ofCOVID-19. The underlying uncertainty distorted mitigation and control efforts around the world (Almadhi et al., 2021). To the extent individual protective behavior reacts to information, unreported cases understate the scale of the epidemic fuelling avoidance of testing and costly self-isolation. The paper argues that test avoidance behavior is less likely to be more pronounced among those without physical disease symptoms, because they tend to discount the immediate threat of infection, at least when observed infection prevalence is low. Such prevalence-dependent T&I decisions, when embedded in a traditional epidemiological model, generate some key insights on the equilibrium dynamic of the epidemic, including a behavioral foundation of why asymptomatic transmission critically drives the spread of SARS-CoV-2.

The results show that the *effective reproduction number* – the key epidemiological measure informing public health policies – is substantially underestimated if asymptomatic infections are disregarded, leading to substantial discounting of the need for stringent pandemic control measures (Arons et al., 2020). Even when the possibility of asymptomatic infections is considered, behavioral barriers facing the asymptomatic agents that fuel propagation of the disease remain unrecognized in health policy approaches so far. The results highlight that undetected asymptomatic infection is a source of downward bias in the COVID-19 fatality rate. Asymptomatic deaths that are only partly accounted for in data therefore potentially explain the number of ‘excess deaths’ reported across the world in the first wave of the pandemic.

The paper stresses the need for policymakers to understand how behavior interacts with reported infection prevalence. At high levels of ‘known’ cases, stringent public policies, such as harsh lockdowns, may be ‘over the top’ because the behavior of the asymptomatics ‘flatten’ the curve automatically.

On the other hand, when detected cases are low, stringency of lockdown measures may be sub-optimally low if the authorities fail to account for low T&I behavior. Indeed, it seems that in the United States, as disease incidence fell after the first wave, costly private prevention efforts declined, prompting relaxation of public disease control efforts, leading to re-emergence of SARS-CoV-2 infections (Atkeson, 2021).

The inherent difficulty in determining the incidence and transmission capability of asymptomatic cases should not lead to underestimating their potent role in the spread of SARS-CoV-2. The findings here emphasize the need to invest in developing a robust global COVID-19 monitoring system, perhaps using the infrastructure developed for worldwide influenza surveillance. Development of a rapid and reliable tracing-and-testing capacity is critical, as suggested by others (Contreras et al., 2021). The simulated model replicating the Italian pandemic scenario in Appendix B suggests strict lockdown measures can be an effective temporary disease containment strategy. Vaccines have presented hopes of long-term protection from SARS-CoV-2, but the recent evidence of ‘breakthrough’ infections in Israel and elsewhere underscore the limitations of vaccine-based approaches to fully eradicate COVID-19 (Dolgin, 2021). The future variants of SARS-CoV-2 may be resistant to the existing vaccines. Hence, even with widespread immunization, it is critical to implement a robust surveillance mechanism that allows for rapid identification of symptomless infections.

## Data Availability

https://www.nature.com/articles/s41591-020-0883-7

## Appendix A

To understand the system behavior, we partition it into 3 subsystems: The first includes just variable *S* (corresponding to susceptible individuals), the second includes *L, I*^*s*^, *I*^*a*^, *T*, (the exposed and infected individuals), which are non-zero only during the transient, and the third includes variables *R* and *D* (representing recovered and dead). We focus on the second subsystem, which we denote the [*L, I*^*s*^, *I*^*a*^, *T*] subsystem.

Variables *R* and *D* (which are monotonically increasing) converge to their asymptotic values 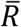 and 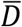, and *S* (which is monotonically decreasing) converges to 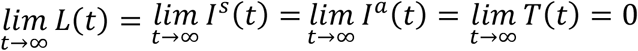.

Define *x* = [*L I*^*s*^ *I*^*a*^ *T*]^*T*^ as the number of individuals in each ‘infected and/or isolated’ compartment. Assume that a Disease-free equilibrium *x*_0_ = {0, 0, 0, 0} exists and is stable in the absence of the disease. We can rewrite the *L-I*^*s*^*-I*^*a*^ *-T* subsystem in the linearized form 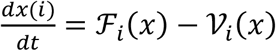 for *i* = 1, …, 4, where ℱ_*i*_(*x*) is the rate of appearance of new infections in compartment *i* and *𝒱*_*i*_(*x*) is the rate of other transitions between compartment *i* and other infected compartments.

Define 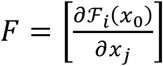 and 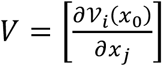 for *i* ≥ 1 and *j ≤* 4.

Biologically, *F* is entry wise non-negative and *V* is a non-singular *M*-matrix, so *V*^−1^ is entry-wise nonnegative. The basic reproduction number is given by: *R*_0_ = *ρ*(*FV*^−1^), where *ρ* denotes the spectral radius. Matrix *FV*^−1^ has (*i, j*) entry equal to the expected number of secondary infections in compartment *i* produced by an infected individual introduced in compartment *j*.

In our [*L I*^*s*^ *I*^*a*^ *T*] sub-model around the initial numbers *S* = *S*_0_, *I*^*s*^ = *I*^*a*^ = *R* = *D* = 0, *T* = *T*_0_, *p*^*a*^(0):

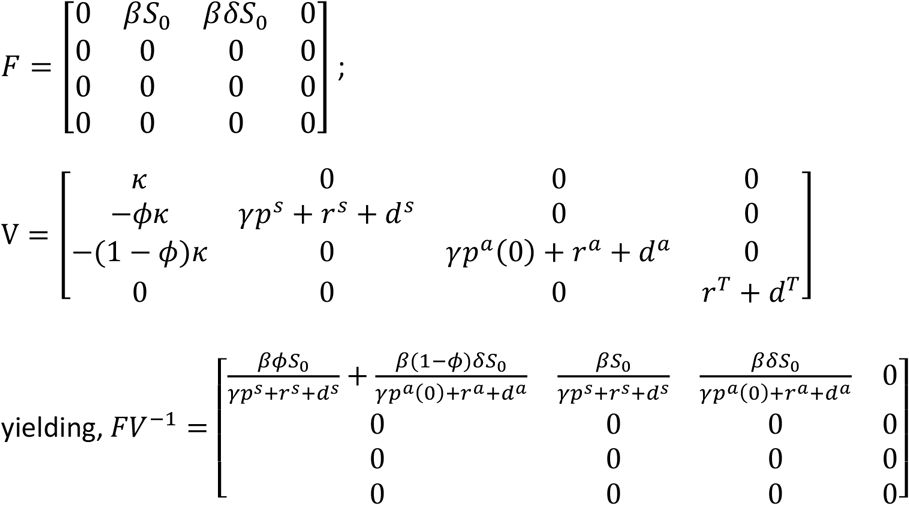

Thus, the behavioral *R*_*0*_ is given by:

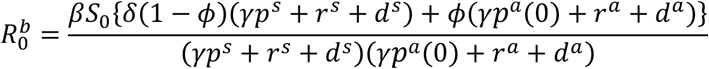

### Derivation of Effective Reproduction Number

In our [*L I*^*s*^ *I*^*a*^ *T*] sub-model around the current numbers *S*(*t*), *I*^*s*^(*t*), *I*^*a*^(*t*), *p*^*a*^(*t*), *T*(*t*), *R*(*t*) and *D*(*t*), and denoting ℵ(*t*) ≡ *S*(*t*) + *T*(*t*) + *R*(*t*) + *D*(*t*), the *M*-matrix is given by:

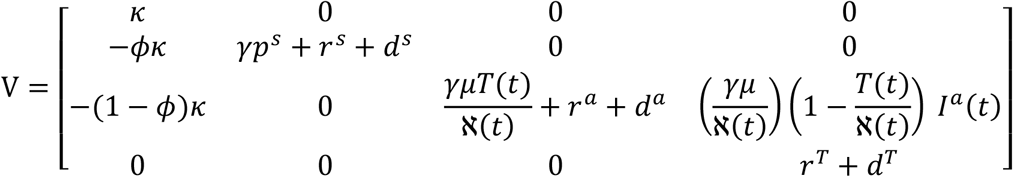

Denoting *ψ*^*s*^ = *r*^*s*^ + *d*^*s*^, *ψ*^*a*^ = *r*^*a*^ + *d*^*a*^ and *ψ*^*T*^ = *r*^*T*^ + *d*^*T*^, we get:

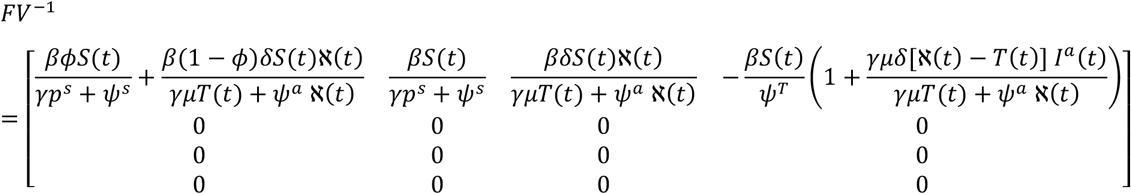

The eigenvalue of this *FV*^−1^ matrix yields the expression in equation (5):

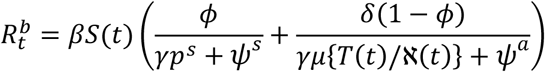

## Appendix B

**Figure B1:**
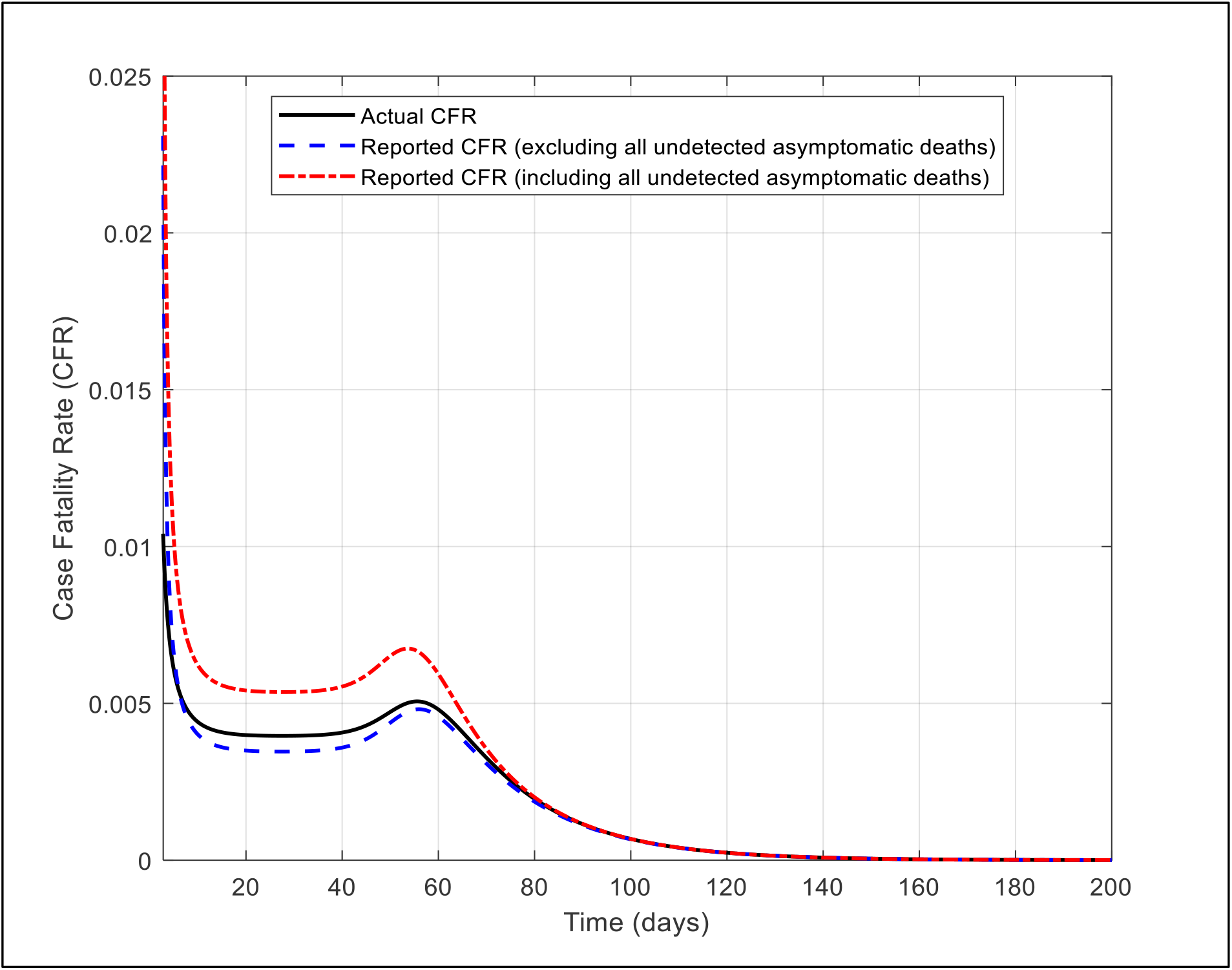
The evolution of actual and reported measures of CFR in the SLIITReD model

**Table B1:**
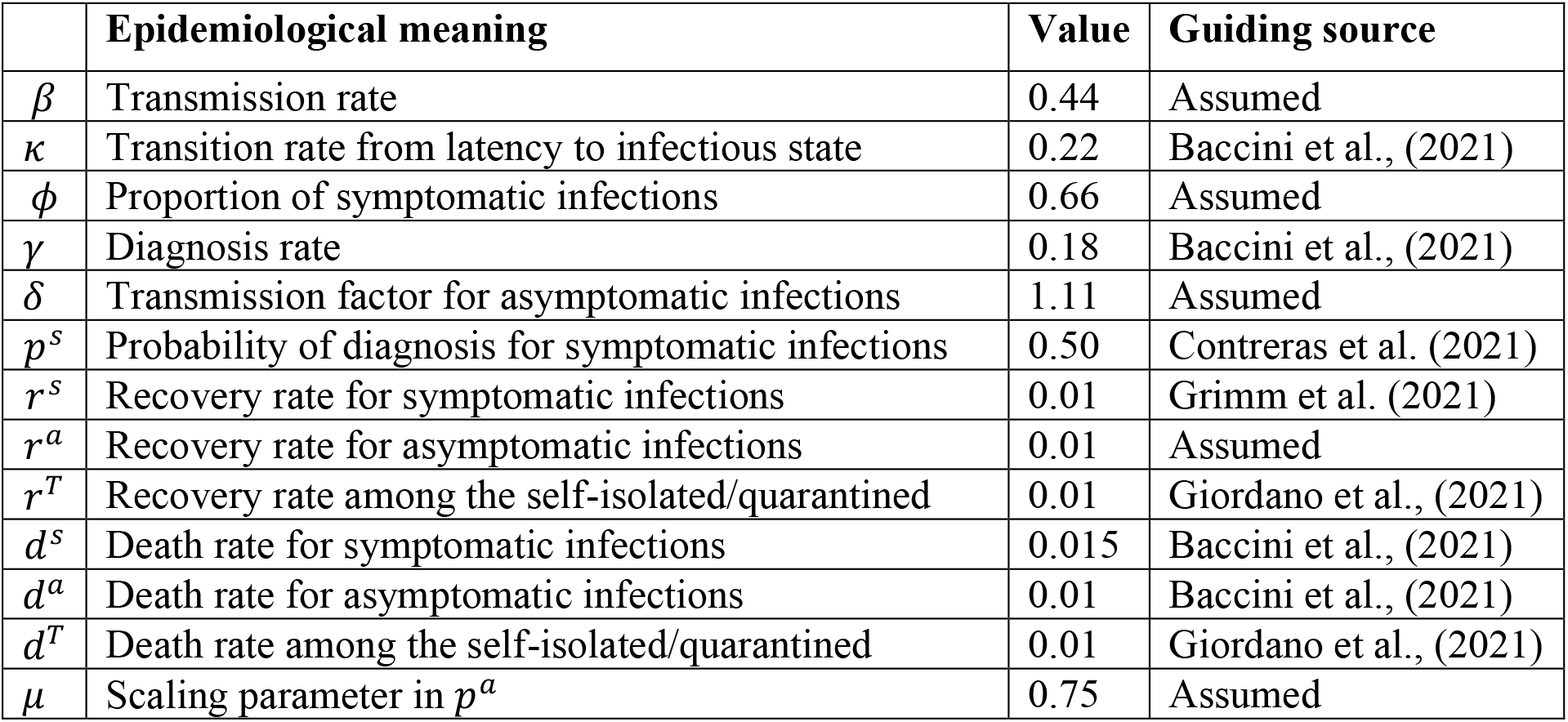
Initial values of the SLIITReD model parameters

|  | Epidemiological meaning | Value | Guiding source |
| --- | --- | --- | --- |
| $\beta$ | Transmission rate | 0.44 | Assumed |
| $\kappa$ | Transition rate from latency to infectious state | 0.22 | Baccini et al., (2021) |
| $\phi$ | Proportion of symptomatic infections | 0.66 | Assumed |
| $\gamma$ | Diagnosis rate | 0.18 | Baccini et al., (2021) |
| $\delta$ | Transmission factor for asymptomatic infections | 1.11 | Assumed |
| $p^s$ | Probability of diagnosis for symptomatic infections | 0.50 | Contreras et al. (2021) |
| $r^s$ | Recovery rate for symptomatic infections | 0.01 | Grimm et al. (2021) |
| $r^a$ | Recovery rate for asymptomatic infections | 0.01 | Assumed |
| $r^T$ | Recovery rate among the self-isolated/quarantined | 0.01 | Giordano et al., (2021) |
| $d^s$ | Death rate for symptomatic infections | 0.015 | Baccini et al., (2021) |
| $d^a$ | Death rate for asymptomatic infections | 0.01 | Baccini et al., (2021) |
| $d^T$ | Death rate among the self-isolated/quarantined | 0.01 | Giordano et al., (2021) |
| $\mu$ | Scaling parameter in $p^a$ | 0.75 | Assumed |

Table B1 describes the parameter values that are inferred from the official data on the evolution of the epidemic in Italy from 20 February 2020 (day 1) through 5 April 2020 (day 46), available at Protezione Civile.^10^ The data horizon (46 days) is chosen to match the work in Giordano et al. (2020), which provides a reliable benchmark.

As in this study, the parameters are updated based on successive public health measures implemented by the policymakers. The fraction of the population in each stage at day 1 is set as: L(1) = 300/60e06, *I*^*s*^(1) = 180/60e6, *I*^*a*^=120/60e06, T(1) = 20/60e06, R(1) = D(1) = 0; *S* = 1 − *L* − *I*^*s*^ – *I*^*a*^ − *R* − *T* − *D*.

After day 4, basic social-distancing measures were implemented and consequently the public became aware of the epidemic outbreak and of the basic hygiene recommendations (such as frequent hand washing, avoiding handshakes and keeping distance) and early school closures by the Italian government, we set *β* = 0.43.

After day 12, a policy came into effect that limited screening to symptomatics only, which decreased the transmission rate to *β* = 0.415 and the probability of testing by the symptomatics from 0.5 to 0.55. People who were completely asymptomatic, did not change their probability of testing.

After day 22, as a result of the partial lockdown *β* = 0.395 and increased the probability of testing by the asymptomatics in response to the rising detected incidence rate.

After day 28, the lockdown became fully operational and stricter (e.g., going out for work was no longer allowed, all non-indispensable activities were stopped gradually). Thus, we set *β* = 0.28, *δ* = 0.66, *γ* = 0.34, and *d*^*T*^ = 0.0012.

After day 38, a wider testing campaign is launched. The parameter values changed to *β* = 0.23, *δ* = 0.29, *γ* = 0.42, and *μ* = 0.79.

The SLIITReD model is simulated using the parameters described above. The behavior of the epidemiological variables predicted by the simulated model are compared with the corresponding values from the official data for the first 46 days are depicted in Figure B2. The trajectories for the current numbers of ‘recovered’, ‘dead’, ‘total infected’, and the ‘number of asymptomatic cases’ are all well-replicated by the SLIITReD model. The simulated model assumes that all deaths of the undiagnosed infected individuals, regardless of their symptom status, are counted in the COVID-19 death tally. Similarly, ‘recovered’ include all recoveries from COVID-19 among symptomatics and asymptomatics. Unlike Giordano et al. (2020), the SLIITReD model reproduces the disease fatality rate well, as shown in Figure B2.

**Figure B2:**
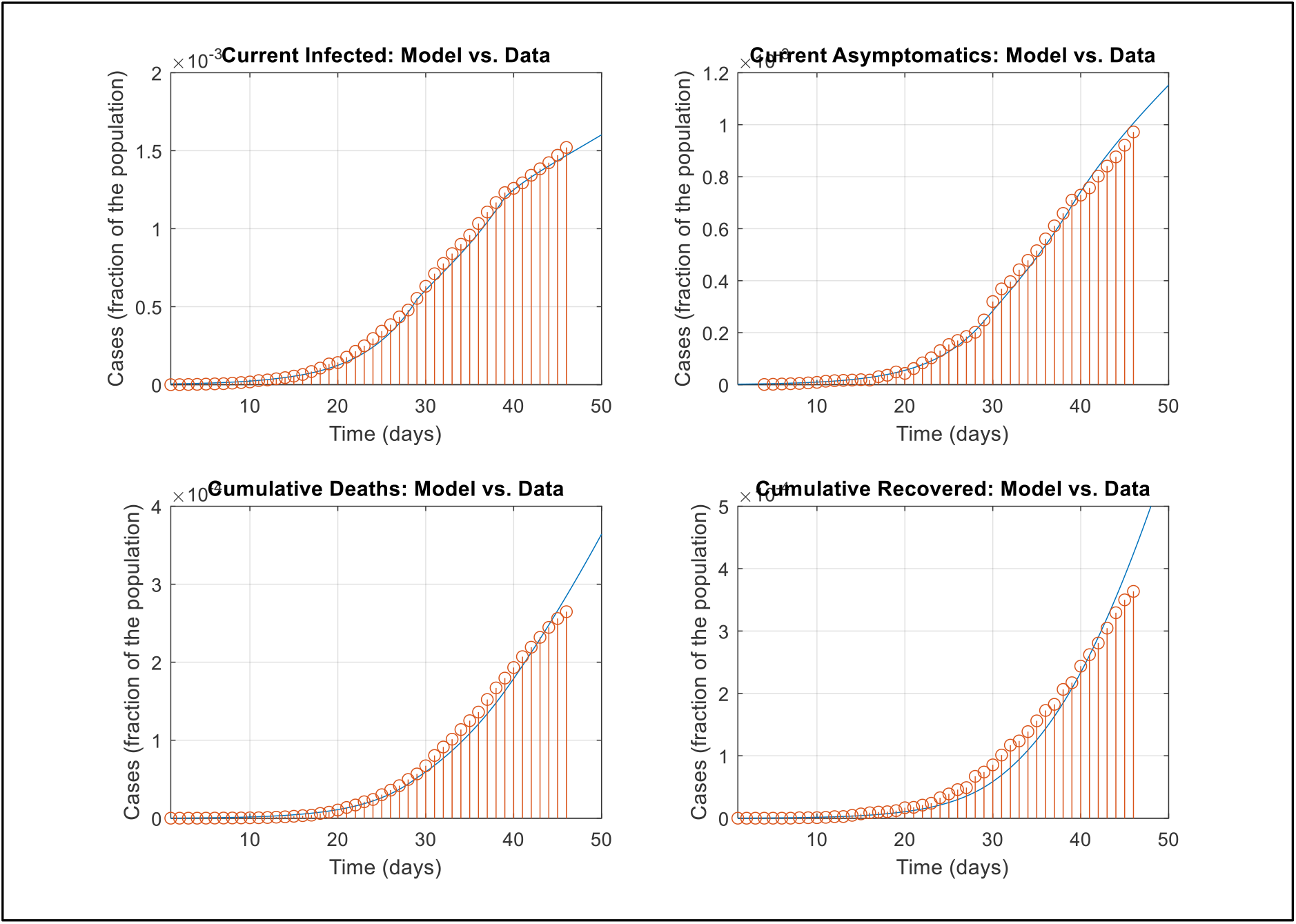
Replication of Italian data by the simulated SLIITReD model

The simulations presented in Giordano et al (2020) were unable to replicate the data on COVID-19 deaths that corresponds to *D*(*t*) in the SLIITReD model – the observed data appeared particularly high with respect to the CFR reported in the literature. In the current simulation, the model is able to reproduce fatality figures. As argued in Giordano et al. (2020), the corresponding CFR is high due largely to the high proportion of older people (50 years and above) in the Italian population and the steep age-gradient of CFR reported across all countries, and by the extensive intergenerational contacts in the Italian society, which propelled the virus transmission from younger to the older and more fragile generations.

The high CFR can also be explained by an overestimation of COVID-19 fatalities in Italy since the official numbers for COVID-19 deaths provisionally included the deaths of all people tested positive for the SARS-CoV-2 virus, even when they had multiple pre-existing comorbidities and the exact cause of death had not yet been ascertained.

While statistical distortion due to provisional data is a challenge in calibrating the model to initial data, in particular with respect to the ratio of fatality to detected cases (CFR), which could be overestimated due to typical over-ascertainment of COVID-19 deaths in the initial phases of a pandemic. However, as discussed in the main text, CFR could also be high due to lower number of detected cases resulting from test avoidance behavior among the asymptomatics when infection prevalence is low.

**Figure B3:**
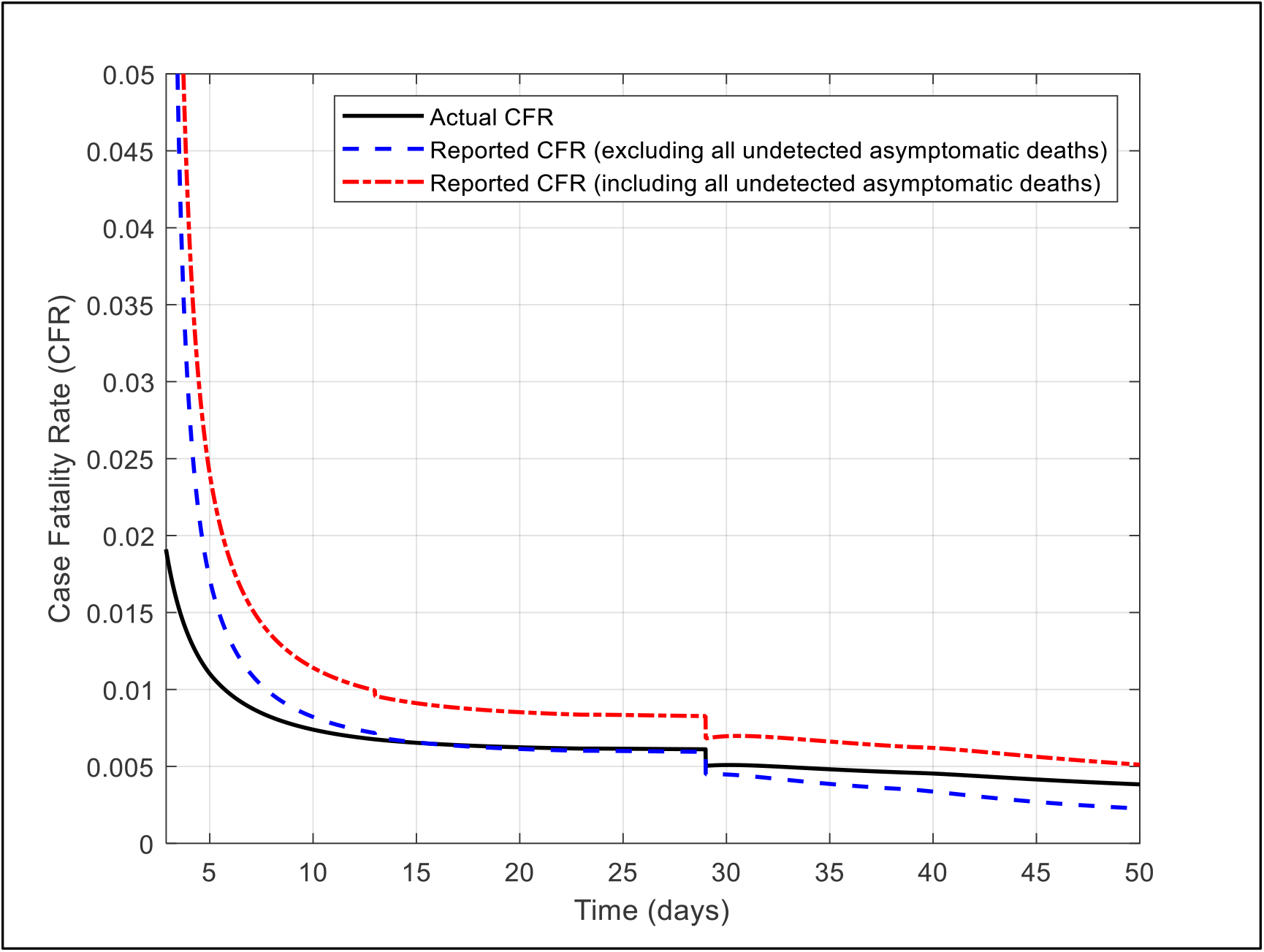
Predicted biases in the COVID-19 fatality measures in Italy

The SLIITReD model reproduced the death figures without substantial fine-tuning of the parameters. Indeed, Figure B3 shows that the reported CFR was higher than the true CFR in the initial phase of the pandemic in Italy. As more cases were detected, the reported CFR converged to the true CFR in the middle phase and fell below the true CFR thereafter, as predicted. Of course, the reported CFR depends on the proportion of deaths among the undetected asymptomatics that are ascertained as COVID-19 deaths. Figure B3 depicts the temporal dynamics of the actual and two reported CFRs – one that excludes and one that includes these deaths. The kink on both curves on day 28 resulted from significant changes in policy measures described above.

The purpose of this replication exercise is not to establish the predictive ability of the model for practice, but simply to show that the model could be a useful guidance to build more accurate predictive models that integrates testing and isolation behavior and the feedback between prevalence and behavior that affects the course of the pandemic itself. The current model has a few caveats as some of the technical assumptions may not be valid in practice. For example, the model implicitly assumes that transition times from one compartment to the next follows an exponential distribution, which may not be true in data. In practice, time spent in a compartment is affected by not only the epidemiological characteristics of the virus, but also by heterogenous social, demographic and economic characteristics across locales and regions affecting the nature of disease propagation (Baccini, 2021).

## Footnotes

1 For example, voluntary use of facial masks during the 2003 SARS epidemic in China, avoidance of visits to public places during the 2009 H1N1 epidemic in Mexico, and self-imposed quarantines of returning US health workers during the 2014 Ebola epidemic.

2 This number was 19 percent at the peak of the first wave in late March 2020. See: https://info.flutracking.net/reports-2/australia-reports/

3 I assume self-quarantine/isolation occurs with full compliance, and self-quarantine/isolation always follows a positive test result. Hence, T&I is considered as a single decision.

4 ‘Excess deaths’ represent the number of people who die from any cause in a given region and period, compared to a historical baseline from recent years. The World health Organization reports an excess death of at least 3 million, as of December 2020. See: https://www.who.int/data/stories/the-true-death-toll-of-covid-19-estimating-global-excess-mortality. A vast proportion of these deaths are directly attributable to undetected SARS-CoV-2 infection (Kowall et al., 2021).

5 As of December 16, 2021, each of the 3158 identified Omicron cases across 27 countries in Europe has been either asymptomatic or mild infections, See: https://www.ecdc.europa.eu/en/news-events/epidemiological-update-omicron-data-16-december. The mild or asymptomatic nature of symptoms could also be due to already existing immunity (vaccine-induced and/or from past exposure) in the infected population.

6 Population can grow, e.g., through new births and inflow of susceptible, infected, and recovered people from outside. Similarly, population can decrease due to death at any of the compartments. These are omitted for simplicity and without loss of generality.

7 Available evidence suggests that test positivity rate (fraction of all COVID-19 tests that return a positive result) closely followed infection rate - see for example, Irons & Raftery (2021).

8 The qualitative results are robust to more general specifications, such as 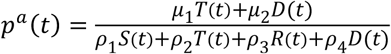 with *μ*_1_, *ρ*_1_ > 0 and *μ*_2_, *ρ*_2_, *ρ*_3_, *ρ*_4_ ≥ 0. The current specification, however, lends itself to a nicer interpretation.

9 The classification system of a COVID-19 death differs across countries. While in some countries, such as the UK, deaths after positive diagnosis of COVID-19 are recorded as COVID-19 deaths, even if the SARS-CoV-2 virus was not the direct cause of death, in other countries COVID-19 deaths may not be preceded by a prior diagnosis of COVID-19.

10 The data is extracted from Giordano et al. (2020), available at: https://www.nature.com/articles/s41591-020-0883-7.

